# Measuring Stigma Associated with Hepatitis B Virus Infection in Sierra Leone: Validation of an Abridged Berger HIV Stigma Scale

**DOI:** 10.1101/2023.02.17.23286086

**Authors:** George A. Yendewa, Edmond J. Sellu, Rashid A. Kpaka, Peter B. James, Sahr A. Yendewa, Peterlyn E. Cummings, Lawrence M. Babawo, Samuel P. Massaquoi, Manal Ghazawi, Ponsiano Ocama, Sulaiman Lakoh, Lawrence S. Babawo, Robert A. Salata

## Abstract

Stigma associated with hepatitis B virus (HBV) is common in endemic countries; however; instruments are lacking to accurately measure HBV-related stigma. We therefore aimed to develop and validate a concise instrument for measuring perceived HBV-related stigma in Sierra Leone. We enrolled 220 people living with HBV (PWHB) aged ≥ 18 years from August to November 2022. The initial Likert-scale instrument entailed 12 items adapted from Berger’s HIV Stigma Scale. We included 4 additional items adapted from the USAID indicators for enacted stigma. The proposed scale’s psychometric properties were assessed. After item reduction, the final HBV Stigma Scale consisted of 10 items and had good internal consistency (overall Cronbach’s α = 0.74), discriminant and construct validity. Exploratory factor analysis produced a 3-dimensional structure accounting for 59.3% of variance: personalized stigma driven by public attitudes (6 items), negative self-image (2 items), and disclosure concerns (2 items). Overall, 72.8% of respondents reported perceived HBV stigma (mean score 29.11 ± 4.14) and a similar a proportion (73.6%) reported at least one instance of enacted stigma. In assessing criterion-related validity, perceived HBV-related stigma correlated strongly with enacted stigma (r = 0.556) and inversely with having family/friends with HBV (r = -0.059). The 10-item HBV Stigma Scale demonstrated good internal consistency and validity and is suitable for screening for HBV-related stigma in Sierra Leone. The psychometric properties of the scale can be optimized with item additions/modifications and confirmatory factor analysis. The scale may help in combating stigma as a barrier to achieving HBV global elimination goals.

## INTRODUCTION

Stigma associated with communicable diseases is widely recognized as an important social determinant of health, and hepatitis B virus (HBV) infection is no exception. Globally, there were an estimated 296 million chronic cases of HBV and 850,000 attributable deaths in 2021 [1]. Sierra Leone and other West African countries are highly endemic (HBV prevalence > 8%) and are among the most adversely impacted by the HBV epidemic [1, 2]. Although there are concerted efforts to achieve the global elimination of HBV by 2030 [3], millions of people living with HBV (PWHB) especially in sub-Saharan Africa (SSA) lack access to quality healthcare and further face the added social burden of stigma and discrimination associated with the disease [3-5].

### Conceptual framework

In 1963, Goffman [6] defined stigma as an *attribute that is deeply discrediting*, which results in being *disqualified from full social acceptance* [6]. Later, in 2001, Link and Phelan [7] proposed a pathway through which stigma operates within power structures. This starts with *stereotyping* and *labelling*, which *separates* the affected individual from the rest of society as the “other” and culminates in *loss of status*, which then invites unfair treatment and discrimination [7]. Additionally, pervasive stigma disrupts many aspects of life in such profound ways as to be considered a fundamental social determinant of health outcomes [8].

Despite the huge global burden of HBV, less is known about the characteristics of stigma associated with HBV, due in part to the lack of validated instruments to accurately measure HBV-related stigma. The existing literature on HBV-related stigma is sparse and comes largely from developed countries, with little representation from SSA [4, 5]. The emerging evidence suggests that HBV-related stigma often arises from knowledge deficits and misconceptions of disease processes [4, 5, 9], although paradoxically, higher levels of knowledge especially of transmission dynamics has been associated with more societal stigma [10, 11]. Additionally, stigma serves as a major barrier to HBV status disclosure. In more collectivist communities, status disclosure can lead to myriad negative social consequences including family and social isolation [12-15], denial of sexual intimacy and in extreme cases partner/spousal abandonment [16-18], and loss of employment opportunities [15, 19]. In the healthcare setting, PWHB may face neglect from providers, which can result in treatment nonadherence and disengagement from care [14, 20-22]. Such negative experiences can cause significant emotional distress such as shame, anger, despair and resignation, resulting in serious mental health problems including anxiety, depression, and post-traumatic stress disorder [18, 23, 24]. Thus, identifying and addressing stigma of the utmost importance, as it has the potential to alleviate social marginalization [4, 5], enhance mental wellbeing and quality of life [22-25], and improve healthcare experiences [26-28].

To design evidence-based interventions that can effectively address HBV-related stigma in endemic countries in SSA, it is crucial to accurately measure stigma levels and its dimensions using culturally-sensitive instruments with robust psychometric properties. Thus, the aim of this study was to develop and validate an HBV Stigma Scale in a cohort of PWHB in Sierra Leone.

## METHODS

### Participants, research sites and recruitment

Study participants were recruited at two hospital-based treatment clinics in Sierra Leone from August to November 2022. Bo Government Hospital (BGH) is a secondary health facility located in Bo, Sierra Leone’s second largest city. The facility is a 400-bed referral hospital in the Southern Province and provides vital services to over 1 million people. Kenema Government Hospital (KGH) is a 350-bed regional hospital serving 670,000 people in Kenema and other districts in the Eastern Province of Sierra Leone.

Potential study participants were approached in treatment clinics at BHG and KGH by trained research staff and informed about purpose of the study. Those who express interest and signed informed consent form were enrolled in the study. The inclusion criteria were: (1) age > 18 years, (2) confirmed HBV infection status by an approved diagnostic method such as serological testing or polymerase chain reaction methods or documentation of infection in clinic records; (3) aware of HBV infection status for ≥ 2 weeks, and (4) able and willing to give informed consent. Exclusion criteria included age <18 years, being unable or unwilling to give informed consent, or not having complicated or terminal illness (e.g., current hospital admission for gastrointestinal bleeding, decompensated liver cirrhosis or hepatocellular carcinoma).

The research staff were from two public tertiary educational institutions in Sierra Leone. Njala University (NU) is located at two campuses in Njala and Bo in southern Sierra Leone. NU is divided into 8 schools and several departments and divisions. The Eastern Technical University (ETU) is Kenema, with several campuses located throughout the Eastern Province. The ETU consists of 2 schools and 4 faculties. Researchers from the School of Community Health Sciences (NU, Bo Campus) and the School of Nursing and Medical Laboratory Sciences (ETU, Kenema Campus) participated in this study. All the research staff who administered the questionnaires to study participants were native Sierra Leoneans with experience in survey methods and intimate knowledge of the local context, including local languages, norms, and customs. A one-week training seminar was conducted prior to the commencement of the study, during which the survey methods and instrument contents were discussed and modified where necessary based on the feedback received from researchers.

### Instrument development and procedures

Survey questionnaire and stigma instrument development was informed by our experiences gained from interactions with PWHB in the clinical and community settings in Sierra Leone, our preliminary work describing the epidemiological, clinical and social aspects of the HBV epidemic in Sierra Leone [11, 29-33] and the broad literature on measuring HIV-related stigma [34, 35], which has been adapted to inform studies in understanding HBV-related stigma [4, 5].

The first section of the survey tool entailed a questionnaire capturing sociodemographic characteristics and HBV-related history of the study participants, i.e., HBV status disclosure, duration since diagnosis, and having family/friends who were PWHB. Furthermore, we included four indicators of enacted stigma, which were adapted from the United States Agency for International Development (USAID)-recommended indicators for assessing discriminatory attitudes towards people living with HIV [36]. The selected indicators assessed (1) partner/spousal abandonment, (2) isolation from family members, (3) social exclusion, and (4) loss of financial resources (workplace stigma) [36]. Responses were recorded as “yes”, indicating that a participant had been the subject of a discriminatory experience, and “no”, indicating the contrary experience. A question answered “yes” earned 1 point, while “no” earned no points, with possible scores ranging 0-4 for enacted HBV stigma.

The second section of the survey tool entailed a 12-item HBV Stigma Scale (Figure 1), which was adapted from Reinius et al [37]. This is an abridged version of the 40-item Berger HIV Stigma Scale originally developed and validated by Berger et al [34]. The full-length Berger scale is one of the most widely used instruments for assessing perceived HIV-related stigma from the perspective of people living with HIV (PWH). It measures stigma across four domains (subscales), as follows: (1) *personalized stigma* (perceived consequences of people knowing about one’s HIV status), (2) *disclosure concerns* (worries about HIV status disclosure), (3) *concern with public attitudes* (accesses discriminatory attitudes from the public), and (4) *negative self-image* (harboring negative feelings towards self internally) [34]. All items are positively keyed and responses are rated on a 4-point Likert scale with equidistant scores, as follows: 1 = strongly disagree, 2 = disagree, 3 = agree, 4 = strongly agree, with higher scores indicating higher levels of perceived stigma. The original Berger HIV Stigma Scale demonstrated excellent psychometric properties with high internal consistency (overall Cronbach’s α = 0.96) and test-retest correlation (r = 0.89–0.92) [34]. The Berger HIV Stigma scale has been validated for use in SSA [38, 39], and full-length or abridged versions have been validated for measuring stigma related to HBV [40], hepatitis C [41] and coronavirus disease 2019 [42].

**Figure 1.**
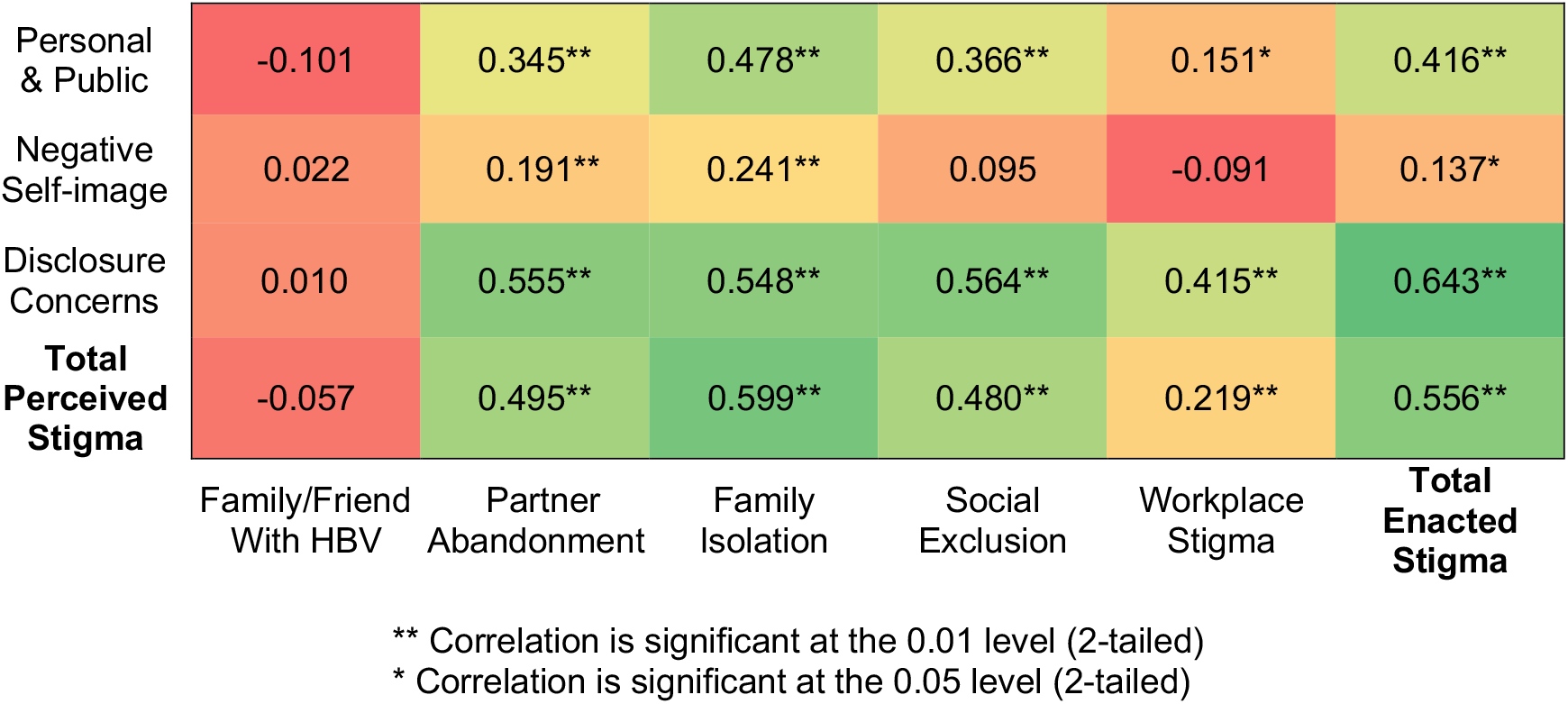
Heatmap of Pearson correlations of the subscales of perceived and enacted HBV stigma

The abridged Berger HIV Stigma Scale by Reinius et al [37] was adapted for this for this study after examining its *content* and *construct validity*. The abridged scale contains 12 items (total score range: 12-40) and replicated the 4-dimensional structure of the original Berger scale, while retaining high internal consistency (Cronbach’s α = 0.80–0.88 across subscales) [37]. In our adaption of the scale, we replaced “HIV” with “Hepatitis B” in all questions and instructions. After slight modifications, the English version was translated by a professional local translation service into the Krio language, the *lingua franca* and *de facto* national language spoken by nearly the entire population of Sierra Leone. The accuracy of the content and cultural appropriateness of the Krio version were verified by the local researchers who are fluent in both English and Krio through back-translation. The Krio version was used to assist participants who could not read or write in English in answering the survey questions. The final English and Krio versions of the questionnaires (Supplementary Materials) were piloted to 10 individuals who were not included in the final study to ascertain the instruments’ clarity and cultural appropriateness. Lastly, to assess *criterion-related validity*, our hypothesis was that PWHB who had experienced at least one instance of discrimination (enacted stigma) would report higher levels of perceived stigma, while having family/friends with HBV would inversely correlate with overall perceived stigma in accordance with Allport’s contact hypothesis [43].

### Statistical analysis

All analyses were performed in SPSS Version 29.0 (Armonk, NY, USA; IBM Corp). Baseline demographic and clinical characteristics were collected. Descriptive statistics was used to summarize baseline sociodemographic and health characteristics. Categorical variables were reported as counts (percentages) and continuous variables were presented as medians (interquartile ranges, IQR).The distribution of stigma scores was accessed by means of the Kolmogorov–Smirnov test of normality and Levene’s test for homoscedasticity, which assumed homogeneity of variances under the null hypothesis.

The psychometric properties of the adapted 12-item HBV Stigma Scale were accessed by estimating its internal consistency (reliability), discriminant validity of items, and construct validity. Internal consistency was demonstrated by means of the following criteria: communality value (h_i_) > 0.20, corrected item-total correlation ≥ 0.32, and coefficient of Cronbach’s α > 0.70. To assess discriminant validity, we calculated the discrimination index (DI) of each item using Kelley’s method [44], which compared the highest (73^rd^ percentile) and lowest (27^th^ percentile) stigma scores, with a DI ≥ 0.20 considered acceptable. We performed exploratory factor analysis using principal axis factoring with orthogonal (Varimax) rotation to assess the dimensional structure of the scale. For factor retention, eigenvalues > 1 and loading saturations > 0.4 were considered. Adequacy of sampling was ascertained using the following conditions: (1) determinant of correlation matrix tending towards 0 (|r|*<* 0.0001), (2) Kaiser-Meyer-Olkin (KMO) index > 0.6, and (3) a statistically significant Bartlett’s Test of Sphericity (BTS). The correlations between factors (subscales) were calculated by Pearson’s correlation coefficient (r). In all analyses, statistical significance for associations were set at *p* < 0.05.

### Ethical approval

Ethical approval was obtained from the Njala University Institutional Review Board (approval date 11 June 2022). Prior to enrolment, it was explained to participants that providing written consent and answering the survey questions implied informed consent.

## RESULTS

### Sociodemographic and Health-related Characteristics of Respondents

We enrolled 220 PWHB in the survey (100% response rate on 12 items), of which 53.6% were from Kenema (Eastern Province) and 46.4% were from Bo (Southern Province). Table 1 describes the baseline sociodemographic characteristics of study participants. The majority were male (54.5%) and the median age was 33 years (IQR 27-41). Nearly half (48.2%) were single and Muslim (49.5%). Most (78.2%) had received primary education or higher. The median duration from HBV diagnosis was 2 years (IQR 1-3) and most ((91.4%) were on treatment. The self-reported HBV disclosure rate was high (72.7%) but fewer than two-fifths (39.5%) reported knowing someone with HBV. Of the USAID-derived indicators of enacted stigma, 66.4% reported workplace stigma, 46.3% had experienced social exclusion, 43.6% reported spousal/partner abandonment and 49.6% felt isolated within their families.

**Table 1.** Baseline characteristics of participants (N=220)

| Characteristics | N (%) |
| --- | --- |
| **Gender** |  |
| Male | 120 (54.5) |
| Female | 100 (45.5) |
| **Age, years** |  |
| Median (IQR) | 33 (27-41) |
| <30 | 80 (35.4) |
| 30-49 | 97 (44.1) |
| ≥50 | 43 (19.5) |
| **Relationship status** |  |
| Single | 107 (48.6) |
| Married | 80 (36.4) |
| Divorced/widowed | 33 (15.0) |
| **Highest education attained** |  |
| None | 48 (21.8) |
| Primary | 25 (11.4) |
| Secondary | 80 (36.4) |
| Tertiary | 67 (30.5) |
| **Religion** |  |
| Christian | 111 (50.5) |
| Muslim | 109 (49.5) |
| **Residence** |  |
| Kenema (Eastern Province) | 118 (53.6) |
| Bo (Southern Province) | 102 (46.4) |
| **Time living diagnosed with HBV** |  |
| Median (IQR), years | 2 (1-3) |
| **Disclosed HBV status** |  |
| Yes | 160 (72.7) |
| No | 60 (27.3) |
| **Has family member or friend with HBV** |  |
| Yes | 86 (39.1) |
| No | 134 (60.9) |
| **Felt abandoned by spouse or partner** |  |
| Yes | 96 (43.6) |
| No | 124 (56.4) |
| **Felt isolated by family members** |  |
| Yes | 87 (39.5) |
| No | 133 (60.5) |
| **Felt excluded from social activities** |  |
| Yes | 103 (46.8) |
| No | 117 (53.2) |
| **Think illness has affected career progression** |  |
| Yes | 147 (66.8) |
| No | 73 (33.2) |

### Descriptive Statistics, Internal Consistency and Discriminant Validity

The internal consistency of all 12 items was first accessed. The overall Cronbach’s α was 0.67. After removing items Q5 and Q12, the internal consistency of the remaining 10 items improved, with overall Cronbach’s α = 0.74. Table 2 shows the descriptive statistics, DI and psychometric properties of the remaining 10 items. The mean stigma score was 29.11 (SD = 4.14). The item mean scores were normally distributed (Z_K-S_ = 0.29, p = 0.200) and the assumption of equal variances (homoscedasticity) was met (F = 5.11, p = 0.330). The coefficients of Cronbach’s α of the individual items on the 10-item HBV Scale varied from 0.67 to 0.74 (Table 2), indicating acceptable reliability of responses. The 10-item scale was retained, as deleting additional items did not improve its overall internal consistency (Cronbach’s α = 0.74). Initial communalities ranged from 0.47 to 0.71 and corrected item-total correlation ranged from 0.21 to 0.63 except for item Q6, which had corrected item-total correlation of 0.08. Items had acceptable discriminant validity (DI range 0.20 to 0.38), except for items Q4 and Q9, which had DI = 0.18 (below the 0.20 threshold). Thus, overall, the 10-item HBV scale demonstrated good psychometric properties and was retained.

**Table 2.** Descriptive statistics, homoscedasticity, discrimination, and internal consistency of adapted HBV stigma scale

| Item | Descriptive Statistics |  |  | Kolmogorov–Smirnov test |  | Homoscedasticity |  | Discrimination |  |  | Internal consistency |  |  |  |
| --- | --- | --- | --- | --- | --- | --- | --- | --- | --- | --- | --- | --- | --- | --- |
|  | Mean ± SD | Skewness | Kurtosis | Z | p | F | p | DI | t | df | p | h<sub>i</sub> | rITC | α<sub>t-i</sub> |
| Q1 | 3.05 ± 0.83 | -0.49 | -0.50 | 0.23 | <0.001 | 8.70 | 0.004 | 0.32 | 10.4 | 95.03 | <0.001 | 0.47 | 0.46 | 0.67 |
| Q2 | 2.56 ± 0.82 | 0.03 | -0.54 | 0.23 | <0.001 | 0.60 | 0.440 | 0.38 | 14.9 | 116.71 | <0.001 | 0.63 | 0.63 | 0.68 |
| Q3 | 2.63 ± 0.95 | -0.26 | -0.83 | 0.25 | <0.001 | 0.61 | 0.437 | 0.38 | 12.6 | 106.99 | <0.001 | 0.71 | 0.50 | 0.67 |
| Q4 | 3.36 ± 0.70 | -1.11 | 1.58 | 0.28 | <0.001 | 3.82 | 0.053 | 0.18 | 2.9 | 106.08 | 0.005 | 0.58 | 0.19 | 0.72 |
| Q6 | 2.67 ± 0.85 | -0.54 | -0.25 | 0.32 | <0.001 | 12.35 | <0.001 | 0.20 | 4.2 | 112.56 | <0.001 | 0.65 | 0.08 | 0.74 |
| Q7 | 3.03 ± 0.75 | -0.57 | 0.26 | 0.29 | <0.001 | 6.24 | 0.014 | 0.22 | 7.1 | 105.40 | <0.001 | 0.62 | 0.34 | 0.69 |
| Q8 | 2.50 ± 0.91 | -0.23 | -0.79 | 0.27 | <0.001 | 1.04 | 0.310 | 0.32 | 9.2 | 116.88 | <0.001 | 0.67 | 0.36 | 0.71 |
| Q9 | 3.43 ± 0.58 | -0.70 | 1.12 | 0.31 | <0.001 | 3.32 | 0.071 | 0.18 | 7.1 | 91.98 | <0.001 | 0.44 | 0.42 | 0.69 |
| Q10 | 2.72 ± 0.75 | -0.76 | 0.48 | 0.36 | <0.001 | 10.11 | 0.002 | 0.21 | 5.6 | 113.22 | <0.001 | 0.61 | 0.30 | 0.70 |
| Q11 | 3.17 ± 0.63 | -0.59 | 1.37 | 0.33 | <0.001 | 4.35 | 0.039 | 0.25 | 9.2 | 100.74 | <0.001 | 0.54 | 0.58 | 0.67 |

*M*, mean; *SD*, standard deviation; *df*, degrees of freedom; *rITC*, correlation between the item and the scale; *h<sub>i</sub>*, communality; $\alpha_{t-i}$ , Cronbach's alpha coefficient after removing the item (significance $\alpha \geq 0.70$ ).

### Construct Validity and Dimensional Structure

Table 3 shows the results of principal axis factoring with Varimax rotation. The estimated a Kaiser-Meyer-Olkin (KMO) index = 0.701, reflecting adequacy of the sample size for the analysis. Bartlett’s Test of Sphericity (BTS) indicated adequacy of the factor extraction model (*χ*^2^ = 512.11, df = 45, p < 0.001). The determinant of the correlation matrix was 0.092, which fulfilled the condition (|r|*<* 0.0001). These results showed that the 10-item scale was appropriate for factor extraction.

**Table 3.**
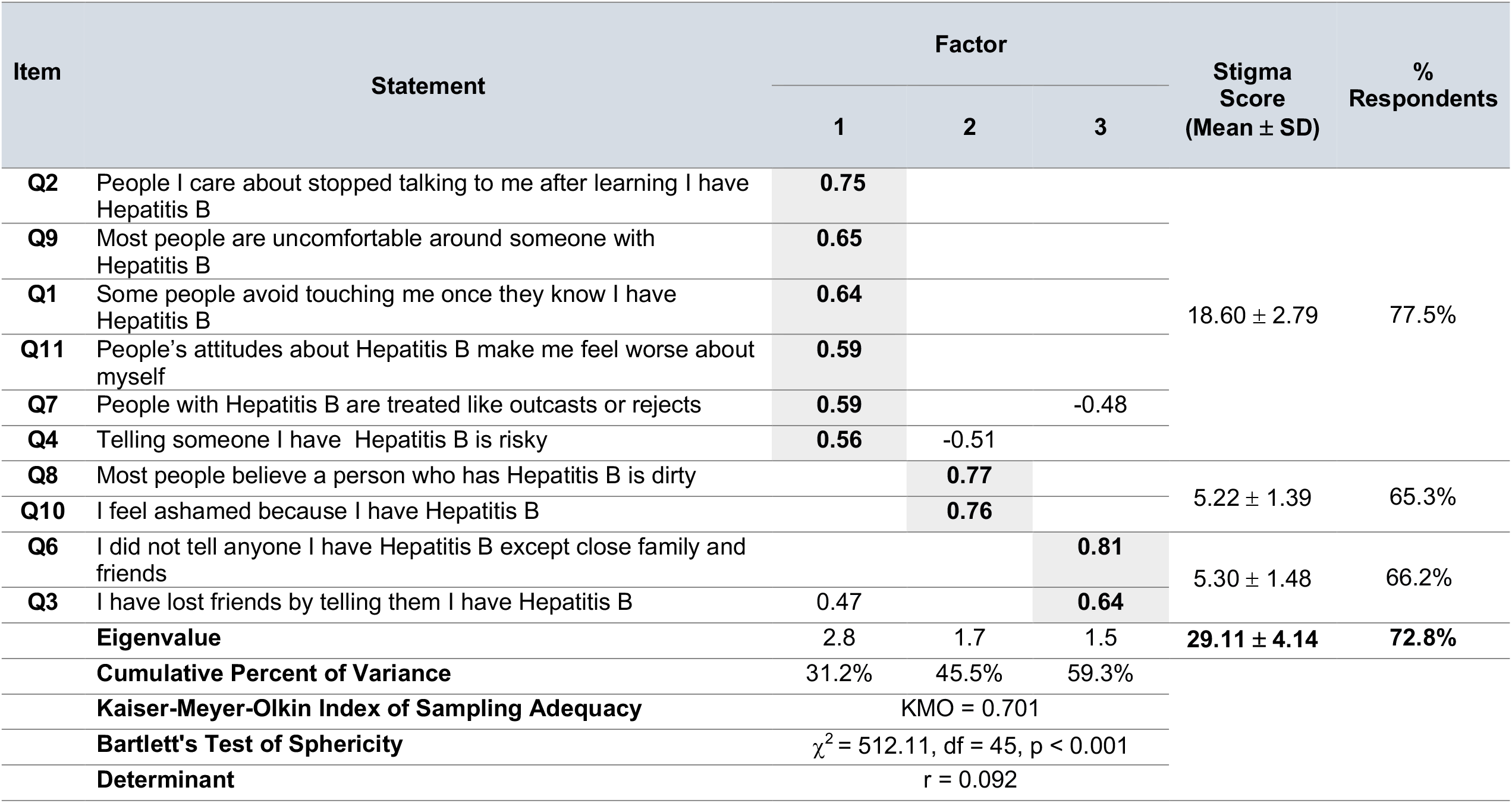
Factor loadings from exploratory factor analysis of HBV Stigma Scale.

The factorial organization of the extracted items consisted of three dimensions which accounted for 59.3% of the total variance (Table 3). The loading saturations were > 0.4. The first factor was comprised of six items (Q2, Q9, Q1, Q11, Q7, Q7 and Q4) and represented “personalized stigma/negative public perceptions”, with subscale Cronbach’s α = 0.74. The second factor consisted of two items (Q8 and Q10) and described “negative self-image”, with Cronbach’s α = 0.66. The third factor integrated two items (Q6 and Q3) and represented HBV “disclosure concerns”, with Cronbach’s α = 0.67. The corrected item-total correlation was high for all items (range 0.36 to 0.63) except for item Q4 which was below the threshold (r = 0.28). These results indicated construct validity of the scale.

### Prevalence of Perceived and Enacted HBV Stigma

The overall mean perceived HBV stigma score was 29.11± 4.14, corresponding to 72.8% of participants (Table 4). Across sub-scales, personalized stigma in response to public attitudes contributed the highest to overall perceived stigma (77.5%, mean score 18.60 ± 2.79), followed by disclosure concerns (66.2%, mean score 5.30 ± 1.48) and negative self-image (65.3%, mean score 5.30 ± 1.48) (Table 4). Based on the USAID-adapted indicators, 73.6% of participants reported at least one instance of enacted stigma. Nearly two-thirds (66.8%) reported having lost career opportunities due to their HBV status. Social exclusion was most frequently reported (46.8%), followed by spousal/partner abandonment (43.6%) and feeling isolated from family members (39.5%).

### Criterion-related Validity and Correlations Between Stigma Subscales

As shown in Figure 1, total perceived stigma correlated strongly with total enacted stigma (r = 0.556, p <0.001). Furthermore, overall perceived stigma correlated highly with three of the four dimensions of enacted stigma: family isolation (r = 0.599, p < 0.001), spousal/partner abandonment (r = 0.495, p < 0.001) and disclosure concerns (r = 0.480). In contrast, overall enacted stigma was highly correlated with two of the three dimensions of perceived stigma: disclosure concerns (r = 0.643, p < 0.001). Loss or resources and negative self-image did not appear to be strongly correlated with overall perceived or enacted stigma. As expected, having family/friend with HBV was inversely correlated with perceived stigma (r = -0.057, p = 0.339).

## DISCUSSION

HBV-related stigma is common in SSA; however, only a small number of studies from the region have described individual and societal perceptions of the disease [11, 12, 14, 20, 21, 45]. Most of these are either qualitative studies which enrolled a small sample size or cross-sectional “knowledge, attitude and practices” studies which incorporated a few items to assess negative attitudes/perceptions of PWHB. Few studies have used validated instruments to measure HBV-related stigma levels. Thus, our study is among the first to develop and validate an instrument for measuring HBV-related stigma in a highly endemic setting in SSA. As such, this study addresses a major gap in the literature. The final HBV Stigma Scale consisted of 10 items and measures stigma from the perspective of PWHB. The scale demonstrated robust psychometric properties, with good internal consistency, content, discriminant and construct validity. An additional advantage of the scale is that because is concise, it can be completed within a few minutes, which makes it suitable for screening for HBV-related stigma in the clinical and community settings.

Exploratory factor analysis of the items revealed a 3-dimensional structure. This was in contrast with the original 12-item abridged HIV Stigma Scale which captures 4 dimensions of HIV-related stigma [37]. The two items each representing the subcategories of “negative self-image” and “status disclosure concerns”, respectively, loaded as distinct factor solutions. Interestingly, however, 6 items representing the remains “personalized stigma” and “stigma due to public attitudes” merged as a unified factor solution. This suggests that personalized stigma may be driven largely by concerns about public attitudes towards PWHB in this setting. The 3-dimenstional structure accounted for 59.3% of variance and the Kaiser-Meyer-Olkin index and Bartlett’s test of sphericity confirmed construct validity of the scale. Although there was cross-loading of three items with high loading saturations (i.e., Q3, Q4, Q7), the items had higher loading saturations and congruity with their parent factors, and therefore did not affect the 3-dimensional structure of the scale. The factorial construct in our study concurs with common themes uncovered from other studies on stigma among PWHB [12-15, 20-24].

The internal consistency of the 10-item HBV Stigma Scale was assessed by three different methods—i.e., communalities, item-total correlation and the coefficient of Cronbach’s α. The overall reliability of the scale was good (Cronbach’s α = 0.74) but lower than the abridged Berger Scale by Reinius et al [37] which was adapted for this study (Cronbach’s α = 0.80–0.88). Across the three subscales, internal consistency was highest for “personalized stigma/public attitudes” (Cronbach’s α = 0.74), while the subscales “negative self-image” and “disclosure concerns” were acceptable but less reliable (Cronbach’s α = 0.66 and 0.67, respectively). Further item reduction did not result in improvement of the coefficient of Cronbach’s α of the two subscales. However, the items fulfilled all other conditions for reliability (all communalities hi > 0.20 and rITC > 0.20), except for items which tested for disclosure concerns, i.e., Q4 (Telling someone I have Hepatitis B is risky) and Q6 (I did not tell anyone I have Hepatitis B except close family and friends). These findings are likely a reflection of cultural differences in how stigma is experienced and perceived [45, 46]. Thus, despite the two items (Q4 and Q6) having low corrected item-total correlations (i.e., rITC < 0.20), they were retained in the final scale owing to their relevance to the conceptual framework. The psychometric indices of the overall scale and in particular the subscales assessing “negative self-image” and “disclosure concerns” can be strengthened by increasing the number of items and/or rephrasing items to improve their discriminant property.

In the present study, 72.8% of PWHB reported perceived stigma and a similarly high proportion (73.6%) reported at least one instance of enacted stigma, confirming a high burden of stigma suffered by PWHB in this setting. Across subscales, personalized stigma/concern with public attitudes contributed the highest to overall perceived stigma (77.5%). Of particular interest was the finding that although status disclosure rates were high (72.7%), nonetheless, nearly two-thirds (66.2%) of participants expressed disclosure concerns. The negative social and metal health consequences of status disclosure have been more extensively studied than other domains of HBV-related stigma. A qualitative study of PWHB from Ghana by Adjei et al [12] showed that fear of stigma and previous negative experiences were consistently expressed as reasons for non-disclosure of status. In a study evaluating the wellbeing of PWHB in China, Li et al [18], HBV status disclosure to sexual partners was associated with significant decrease in all four domains of the World Health Organization Quality of Life assessment (i.e., physical, psychological, social, and environmental). However, the study from Ghana by Adjei et al [12] noted that despite fear of repercussions, status disclosure was also perceived as beneficial in enhancing HBV testing, prevention (vaccination), building trust in relationships and obtaining social/financial support. Similar healthcare benefits from status disclosure (i.e., increased likelihood of HBV testing) have been reported by Franklin et al [21] in Zambia.

Furthermore, our analysis showed that the scale met the conditions for criterion-related validity. Total perceived stigma and its subscales correlated well with total enacted stigma and its components, with the correlation strongest for disclosure concerns (r = 0.415 to 0.643) (Figure 1). Similarly, PWHB reporting having disclosed HBV their status correlated highly with the subscale of stigma associated with disclosure concerns (r = 0.605) and total enacted stigma and its subscales (r = 0.327 to 0.464). On the other hand, interpersonal relationships are an important coping mechanism and the support of family and friends is crucial in dispelling stigma and prejudice related to disease. This was tested with the statement “Do you have a family member, close friend or colleague who has Hepatitis B?”. As expected, having family/friends with HBV had an inverse relationship with overall perceived stigma (r = -0.057), underscoring the importance of family and other social support networks in designing interventions seeking to reduce stigma as a barrier to elimination efforts.

Our study had some limitations worthy of discussing. Firstly, this was a cross-sectional study rather than a mixed-methods approach which would have offered some insights into causal relationships. Secondly, this was a healthcare-based study from two geographic regions in Sierra Leone; thus, its generalizability may be limited. Thirdly, although the overall internal consistency of the scale was good, the reliability of the subscales on “negative self-image” and “disclosure concerns” should be approached with caution and can be improved. This can be achieved by increasing the total number of items in each subscale and/or modifications to strengthen their discriminant property and cultural sensitivity. Additionally, we were unable to perform confirmatory factor analysis due to the small sample size. Finally, lack of other validated HBV stigma instruments from SSA did not allow for comparison of our findings with that of others. Nonetheless, our analysis is among the first to develop and validate an instrument with good psychometric properties for measuring stigma from the perspective of PWHB and will contribute to efforts seeking to combat stigma as a major barrier to HBV elimination in SSA.

## CONCLUSION

In summary, the 10-item HBV Stigma Scale demonstrated good internal consistency and construct validity and is suitable for measuring stigma from the perspective of PWHB. The scale had three dimensions including “personalized stigma driven by concerns about public attitudes” (6 items), “negative self-image” (2 items) and “disclosure concerns” (2 items). The psychometric properties can be optimized with item additions/modifications and further studies are suggested for confirmatory factor analysis. A high proportion of PWHB (72.8%) reported perceived stigma, with a similarly high proportion (73.6%) reporting at least one instance of enacted stigma, confirming a high burden of stigma suffered by PWHB in this setting. This scale is suitable for use in both the clinical and community setting and will help to combat stigma as a major barrier to HBV elimination in SSA.

## Data Availability

All data produced in the present study are available upon reasonable request to the authors

## AUTHOR CONTRIBUTIONS

GAY, LSB and RAS conceptualized and designed the study. EJS, RAK, PBJ, SAY, PEC, LMB, SPM and MG contributed to study concept and design. EJS, RAS, LSB collected the data. GAY conducted the statistical analysis. BBJ, PO and SL contributed important intellectual content. All authors contributed to the interpretation of data. GAY wrote the initial manuscript draft. All authors critically revised and approved of the final version. GAY is acting as the guarantor of this manuscript.

## ACKLOWLEDGEMENTS

We wish to acknowledge the staff at Bo and Kenema Government Hospitals and people living with hepatitis B without whose help this study would not have been successful.

## DATA AVAIL ABILITY STATEMENT

The data presented in this study are available on request from the corresponding author upon reasonable request.

## ETHICAL APPROVAL

Ethical approval was obtained from the Njala University Institutional Review Board (approval date 11 June 2022).

